# Neurocomputational modelling for characterizing neurocognitive pathophysiology in clinical high risk for psychosis

**DOI:** 10.1101/2025.09.16.25335903

**Authors:** Andreea O. Diaconescu, Colleen E. Charlton, Zheng Wang, Milad Soltanzadeh, Jennifer R. Lepock, Alban E. Voppel, Lena K. Palaniyappan, John D. Griffiths, Michael Kiang

## Abstract

**Background:** The N400 semantic-priming event-related potential (ERP) is attenuated in schizophrenia and in youth at clinical high risk for psychosis (CHRP); however, the circuit mechanisms linking this abnormality to functional outcome remain unclear.

**Methods:** We recorded 32-channel EEG while 46 CHRP outpatients and 38 demographically matched healthy controls (HC) performed a word-pair priming task (80 related, 80 unrelated pairs; prime-to-target stimulus-onset asynchronies [SOA]=300ms or 750ms). Twenty-six CHRP participants were reassessed after one year. N400 difference waves (unrelated–related) were fit with a connectome-constrained Jansen–Rit neural-mass model in 200 cortical parcels. Local gains, synaptic time constants and effective connectivity parameters were optimised with the WhoBPyt framework, and principal-component trajectories of the inferred excitatory–inhibitory |*E* − *I*| balance were analysed with partial least squares.

**Results:** Under the long-SOA (750ms)/unrelated condition, CHRP showed a sharply elevated early |*E* −*I*| peak at 70–100ms relative to HC (p=0.0004), driven by greater pyramidal excitatory gain (parameter A), stronger excitatory-to-pyramidal coupling and faster inhibitory decay, indicating cortical disinhibition. The amplitude of this early peak predicted poorer social functioning at one-year follow-up (r=–0.56,p=0.003). Conversely, in the short-SOA (300ms)/related condition CHRP exhibited a larger N400-window |*E*−*I*| peak (350ms) associated with enhanced inhibitory-to-pyramidal feedback (parameter C3) and lengthened inhibitory decay (parameter b); this putative compensatory inhibition correlated with better functional outcome (r=0.60,p=0.001). Network-level analyses revealed an amplified early sensory-network burst and attenuated default-mode and salience-network responses, consistent with a systems-wide shift toward disinhibition in CHRP.

**Conclusions:** Computational modeling demonstrates that N400 abnormalities in CHRP arise from temporally specific E–I imbalances: early cortical disinhibition that forecasts functional decline and a later inhibitory reinforcement that may confer resilience. These time-resolved E–I metrics constitute low-burden, mechanistically interpretable biomarkers for stratifying psychosis risk and guiding early intervention.

## 1. Introduction

Schizophrenia-spectrum disorders affect about 1% of the population [1], often manifesting in young adulthood and causing significant illness burden. For instance, schizophrenia ranks seventh globally for years lived with disability and third among individuals aged 15-44 [2]. The total cost of schizophrenia in Canada is around $7 billion annually [3]. Thus, it is vital to identify young adults at risk of schizophrenia and provide timely and necessary mental health care.

Those exhibiting attenuated psychotic symptoms without meeting the criteria for a psychotic disorder, termed “prodromal” or at clinical high risk for psychosis (CHRP), are of particular interest in this endeavour. Around 3% of young adults seeking mental health care meet CHRP criteria and are at higher risk for psychotic disorders [4]. Notably, current interventions, whether pharmacological or psychological, can significantly improve outcomes in these individuals [5].

Although not all CHRP individuals develop psychosis, the majority of CHRP individuals continue to exhibit persistent symptoms [6, 7, 8, 9]. In contrast, about 30% recover both clinically and functionally over several years [6, 10]. Given these varied trajectories, there is a pressing need to identify better predictors for CHRP outcomes, in order to prioritize treatment interventions. Current clinical variables have limited predictive value [11], and while neuropsychological and neuroimaging measures have shown potential, they can be time-consuming and expensive [12, 13, 14].

Electrophysiological techniques, particularly scalp-recorded ERPs, have been proposed as powerful tools for improving the prediction of outcomes in CHRP [15]. ERPs measure voltage changes corresponding to cognitive events, reflecting synchronous activity of cortical pyramidal neurons. These tools are objective, rater-independent, and more economical than some alternatives such as functional magnetic resonance imaging (fMRI) [15].

A well-established ERP measure, the N400 waveform, holds potential for psychosis prediction in CHRP individuals [16]. This negative-going voltage emerges approximately 400 ms after a semantic stimulus like a word or image and is prominent across the scalp, reflecting activity in the inferior and anterior medial temporal lobe [17]. These “N400 semantic priming effects” indicate the contextual prediction of upcoming items by activating associated neural concepts in our long-term semantic memory [18]. Essentially, the N400 serves as a semantic domain’s prediction error index [18].

The N400 has been employed to study atypical semantic processing in schizophrenia. Many studies have identified aberrant N400s in schizophrenia [19, 20, 21, 22, 23], especially with a stimulus-onset asynchrony (SOA) of “prime” and “target” words of at least 400 ms or more [24]. This reflects schizophrenia individuals’ difficulty in using context to activate related concepts in semantic memory [25]. There is growing evidence that N400 semantic priming deficits could be a psychosis biomarker [26, 21], as they have been linked to delusions and paranoid ideation in non-clinical samples [27, 28]. These deficits have been observed in CHRP individuals, particularly with longer SOAs [29, 30], indicating similar challenges in controlled semantic priming as in schizophrenia patients [21, 31].

We have recently shown that N400 semantic priming deficits can predict one-year clinical outcomes in CHRP [32, 30]. For instance, reduced N400 semantic priming at a 750-ms SOA is associated with declining social functioning one year later [30]. However, this predictive utility did not hold over two years [32], indicating that N400 measures may be short-term rather than long-term predictors of social functioning. These outcomes align with other studies where atypical semantic processing, such as disconnectedness in transcribed speech samples in CHRP individuals correlated with increased psychosis onset and poorer functioning, evidenced by higher conversion rates to psychosis at both one [33] and seven-year marks [34]. Additionally, CHRP showing atypical fMRI activation during semantic tasks had poorer symptomatic and functional outcomes within 6-24 months [35]. Additional research is needed to confirm the prognostic relevance of the N400 ERP marker and evaluate if it can enhance long-term prediction accuracy.

In our study, we aim to leverage neurocomputational modeling to assess the neural mechanisms explaining the abnormalities in N400 indices in CHRP. The N400, dependent on NMDA receptor (NMDAR) plasticity [36], and its reduction in schizophrenia suggest NMDAR hypofunction as a potential mechanism of functional decline in CHRP [37]. This mechanism may underlie the perceptual and linguistic disturbances observed in schizophrenia and CHRP, akin to the effects of the NMDAR antagonist ketamine in healthy individuals [38, 39].

By utilizing neurocomputational models to analyze N400 data from CHRP individuals, we aim to test the first hypothesis that the reduced N400 priming effects in CHRP compared to controls stem from reduced NMDAR plasticity and disruptions in pyramidal cell excitation of inhibitory interneurons via glutamatergic NMDA receptors. Deficiencies in the structure and function of glutamate synapses are fundamental to the neurobiology of schizophrenia and may represent some of the earliest pathological changes associated with the disorder [37]. Genetic research has highlighted that both rare [40, 41, 42] and common [43, 44] gene variants associated with schizophrenia involve those responsible for the development, functioning, and elimination of glutamate synapses. These critical genes are particularly active in the frontal cortex during prenatal development and shortly after birth [45, 46]. Specifically, deficits in NMDAR-mediated glutamate signaling, especially in layer 3 pyramidal neurons of the prefrontal cortex [47], are believed to contribute to impaired executive functions such as working memory [48].

The proposal of NMDAR hypofunction in schizophrenia is also supported by the psychotomimetic effects of ketamine in healthy subjects [49, 50, 51] and its exacerbation of psychosis symptoms in those with schizophrenia [52]. Additionally, the implicated role of GABAergic dysfunction points to an intertwined pathology involving NMDARs and inhibitory interneurons [53]. We predict that this neurobiological disruption could predict more severe symptoms and poorer 1-year functional outcomes in CHRP individuals.

To explore these hypotheses, we employ a connectome-based neural mass modelling (CNMM) where neural dynamics at each source are described by Jansen-Rit (JR) equations [54, 55]. The JR model is a widely used neurophysiological model for both stimulus-evoked and resting-state EEG measures [56] and has been previously been employed to examine reduced MMN effects in schizophrenia patients [57, 58, 59, 60]. This neural mass model encapsulates neural dynamics across three populations: pyramidal neurons, excitatory, and inhibitory interneurons, forming a circuit with one positive and one negative feedback loop, see Figure (1).

**Figure 1:**
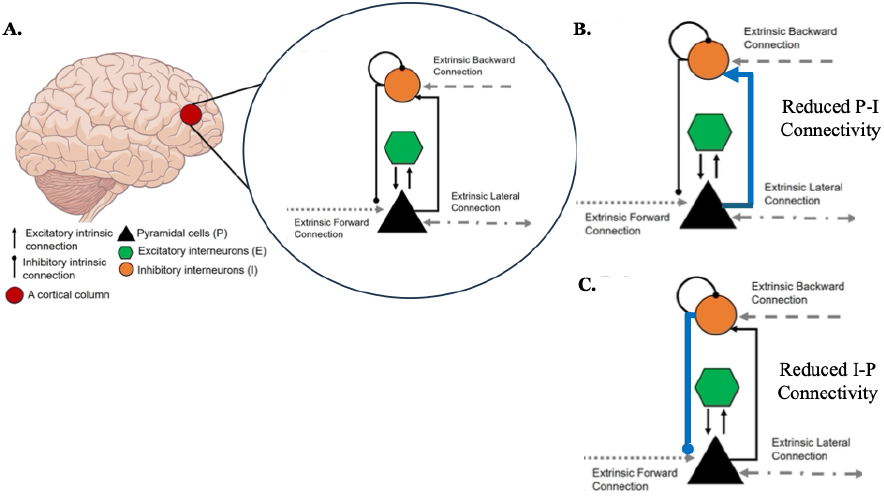
Hypothesized neurobiological mechanisms underlying N400 abnormalities in individuals at clinical high-risk for psychosis (CHRP): By utilizing a connectome-based neural mass modeling approach, we aim to elucidate the neural circuitry disruptions that may predict more severe symptoms and poorer functional outcomes in CHRP. This figure schematic illustrates the proposed circuit involving pyramidal neurons and interneurons, highlighting the feedback loops critical to understanding the neurobiological disruptions in psychosis.

## 2. Methods

### 2.1. Participants

We enrolled 46 CHRP participants and 38 HC participants, matched for age and premorbid IQ, as estimated by the National Adult Reading Test [61]. ERP data from some of these participants were presented in prior studies [29, 30]. CHRP participants were help-seeking outpatients referred to the Clinical High Risk (CHRP) Clinic at the Centre for Addiction and Mental Health (CAMH), Toronto. HC participants were recruited from the community through online advertisements, newspapers, and bulletin boards. All participants provided written informed consent, and the study received approval from the CAMH Research Ethics Board.

CHRP individuals met the diagnostic criteria for a psychosis-risk syndrome according to the Criteria of Psychosis-Risk States, assessed via the Structured Interview for Psychosis-Risk Syndromes (SIPS) [62]. All CHRP participants met criteria for the attenuated psychosis syndrome subtype [63] and had no history of current or lifetime DSM-IV-TR Axis I psychotic disorder, or mood disorder with psychotic features, as determined by the Structured Clinical Interview for DSM-IV-TR. Participants had no history of DSM-IV substance use or dependence in the past six months (with the exception of nicotine) and were antipsychotic-naive. At the two-year follow-up, three CHR individuals had converted to psychosis.

### 2.2. N400 Paradigm and Cloze Probability Analysis

The stimuli used in this study were identical to those employed by [64] and consisted of 80 related (e.g., METAL-STEEL) and 80 unrelated (e.g., DONKEY-PURSE) prime-target word pairs. For each related pair, the target was selected based on the most common associative response to the prime, as indicated by participants in the University of South Florida word-association norms [65]. The mean response probability for related targets—i.e., the proportion of individuals who produced the target word in response to the prime was 0.61 (SD = 0.12). In the case of unrelated pairs, the prime and target were not associated in the norms. Targets were matched across conditions for mean length and log-transformed frequency, and the primes were similarly matched on these parameters. Additionally, 160 pairs comprising a word prime and a pronounceable non-word target (e.g., DRESS-ZORES) were included. No word appeared more than once among the stimuli.

#### 2.2.1. Semantic similarity

We also examined the role of cloze probability, an index of semantic predictability of the stimuli pairs, to assess its relationship with N400 amplitudes and neurocomputational parameters. Using GloVe 6B cosine similarity distance scores (1 to 1), we computed cloze semantic probabilities similarities for each word pair. Related word pairs (mean = 0.603) were more semantically similar than unrelated pairs (mean = 0.163).

The stimulus list was presented in a fixed randomized order across four blocks of 80 trials each. Two sets of stimulus onset asynchronies (SOAs) - short (300 ms) and long (750 ms) SOA conditions - were used to distinguish between automatic and controlled semantic processing, providing insight into cognitive deficits in CHR individuals (Figure 2. Short SOA conditions primarily reflect rapid, automatic lexical activation, relying on associative memory, whereas long SOA conditions require controlled, context-dependent semantic integration, engaging executive functions for predictive processing. Prior research has shown that CHR individuals exhibit impaired N400 semantic priming at long SOA but not short SOA, suggesting deficits in maintaining and integrating context over time[30].

**Figure 2:**
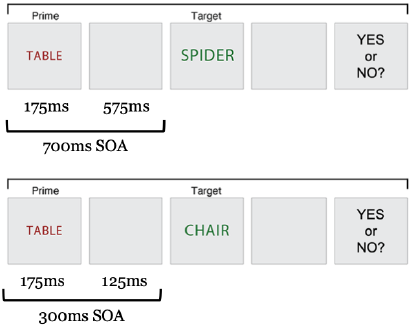
N400 semantic priming paradigm: Participants were presented with 80 related (e.g., METAL-STEEL) and 80 unrelated (e.g., DONKEY-PURSE) prime-target word pairs, along with 160 word-pronounceable non-word pairs (e.g., DRESS-ZORES). Related targets were selected based on common associative responses in established word-association norms, with a mean response probability of 0.61 (SD = 0.12). Unrelated pairs had no such association. Targets and primes were matched for length and frequency across conditions. The stimulus list, presented in four blocks of 80 trials each, was counterbalanced across two versions. Each trial began with a fixation cross (500 ms), followed by a prime word (175 ms), an SOA-dependent blank screen (125 ms or 575 ms), a target word (250 ms), and a prompt (“Yes or No?”) for participants to indicate whether the prime and target were related. Responses were made via button press, and the button assignment was counterbalanced among participants. The trial concluded with a blank screen (3000 ms) before the onset of the next trial.

Each participant viewed the stimuli on a video monitor. Each trial consisted of the following sequence: (a) a preparatory fixation cross displayed for 500 ms; (b) a blank screen for 250 ms; (c) the prime word for 175 ms; (d) a blank screen for 125 ms (in 300-ms SOA trials) or 575 ms (in 750-ms SOA trials); (e) the target word for 250 ms; (f) a blank screen for 1250 ms; (g) a prompt “Yes or No?” displayed until the participant responded via button press; and (h) a blank screen for 3000 ms before the onset of the next trial.

At the prompt, participants were instructed to press one of two buttons positioned under their right and left thumbs. They pressed a button to indicate whether the second word was a real word (“yes”) or a non-word (“no”). The assignment of buttons was counterbalanced among participants and across the two versions of the stimulus list.

### 2.3. EEG Recording and Preprocessing

Continuous EEG data were recorded using an actiCHamp amplifier (Brain Products, Gilching, Germany) from 32 active Ag/AgCl electrodes embedded in a cap (actiCAP system, Brain Products). The electrodes were positioned approximately evenly across the scalp according to a modified International 10–20 System (Fp1, Fp2, F7, F3, Fz, F4, F8, FC5, FC1, FC2, FC6, T7, C3, Cz, C4, T8, TP9, CP5, CP1, CP2, CP6, TP10, P7, P3, Pz, P4, P8, PO9, O1, Oz, O2, PO10). Electrode impedances were maintained below 25 *k*Ω. The EEG was referenced online to FCz and continuously digitized at a sampling rate of 500 Hz. Blinks and eye movements were monitored using electrodes placed on the supraorbital and infraorbital ridges and at the outer canthi of both eyes.

Offline, the EEG data were re-referenced to the algebraic mean of the mastoids and bandpassed at 0.25–60 Hz. Eyeblink artifacts were corrected using principal component analysis according to [66]. Individual trials with artifacts due to eye movement, excessive muscle activity, or amplifier blocking were additionally manually rejected through visual inspection prior to time-domain averaging.

Event-related potentials (ERPs) were computed for epochs ranging from 100 ms before stimulus onset to 900 ms after stimulus onset. For each participant, separate ERP averages were calculated for trials with related and unrelated targets at each SOA. The N400 semantic priming effect was defined as the mean voltage of the difference wave, obtained by subtracting the average ERP for related trials from that of unrelated trials, measured from 300 to 500 ms post-stimulus onset, consistent with previous methodologies [27].

### 2.4. Whole Brain Modeling Framework

The fitting of the Jansen-Rit (JR) model to ERPs across the whole brain using a recurrent neural network (RNN) deep learning framework, specifically implemented in PyTorch, represents a novel approach to understanding the neural dynamics underlying cognitive processes such as semantic processing [67]. The JR model, which simulates the activity of excitatory pyramidal cells and inhibitory interneurons, serves as a foundational framework for capturing the complex interactions that contribute to the generation of ERPs like the N400 waveform, see Figure 3.

**Figure 3:**
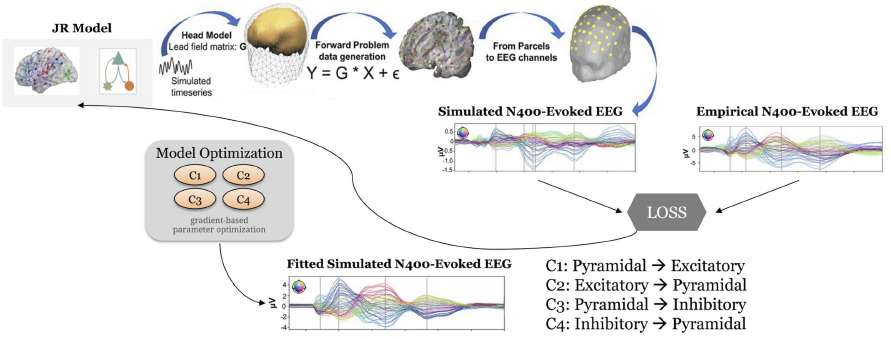
Connectome Neural Mass Model for fitting N400 event-related potentials (ERPs): The Jansen-Rit (JR) neural model is used to simulate the activity of excitatory pyramidal cells, excitatory spiny stellate cells, and inhibitory interneurons, capturing their dynamic interactions through a set of differential equations. The neural time series are transformed via a forward model and lead field matrix to EEG sensor space, generating simulated N400-ERP responses. The model is fit to the empirical N400 ERPs using the WhoBPyt machine learning framework to optimize model parameters via gradient-based backpropagation through time (BPTT), minimizing the loss between simulated and empirical ERPs. This process enables the estimation of neurocomputational mechanisms underlying semantic processing in terms of local gain and effective connectivity parameters and variations in excitatory-inhibitory |*E* − *I*| balance at the individual subject level.

**Figure 4:**
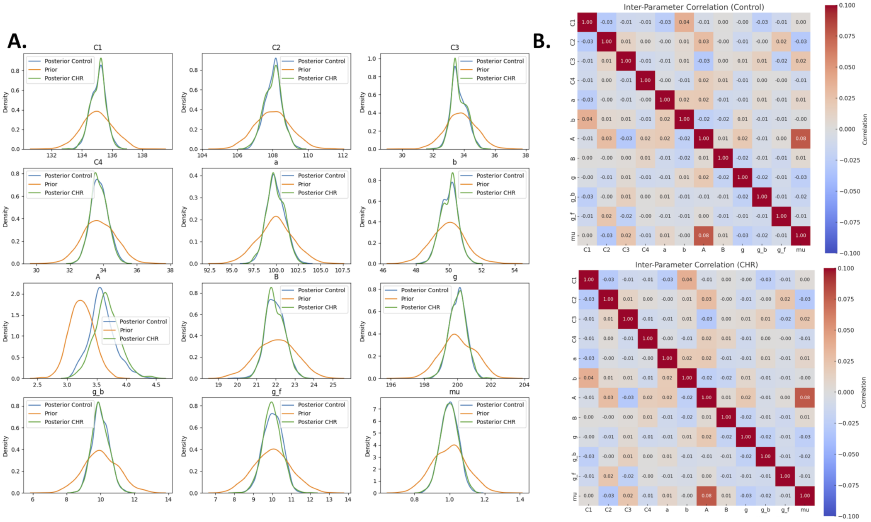
Parameter identifiability of the Jansen-Rit model. **Left (A):** Prior-to-posterior contraction for all fitted parameters (synaptic gains, time constants, and effective couplings; e.g., *A, a, b, C*_1_–*C*_3_). Orange curves show the weakly informative priors; green curves show posterior marginals estimated from the data, plotted separately for healthy controls (HC) and clinical high-risk participants (CHR-P). Posteriors are consistently narrower than priors, indicating information gain from the ERP data. The most pronounced contraction and group-separable shifts are evident for pyramidal gain *A*, inhibitory decay *b*, and inhibitory-to-pyramidal feedback *C*_3_, consistent with a CHR-P bias toward cortical disinhibition. **Right (B):** Posterior inter-parameter correlation matrices (common colour scale [− 0.10, 0.10]) for HC (top) and CHR-P (bottom). Off-diagonal elements are uniformly small (mean |*r*| *<* 0.03, maximum |*r*| *≈* 0.08), indicating minimal multicollinearity and limited parameter trade-offs. Together, the posterior contraction and near-diagonal correlation structure support practical identifiability of the model parameters used in the main analyses.

The Jansen-Rit model consists of three interconnected neural populations: excitatory pyramidal cells, excitatory spiny stellate cells, and inhibitory interneurons [56]. The dynamics of these populations are described by a set of differential equations that capture their excitatory and inhibitory interactions. This model is particularly useful for simulating the macroscopic electrophysiological activity recorded in EEG and MEG studies, allowing the examination of how variations in excitatory-inhibitory |*E* − *I*| balance affect cognitive functions and associated ERPs. To fit the Jansen-Rit model to ERP data across the whole brain, the following steps can be undertaken using the PyTorch framework: Implement the Jansen-Rit model equations within a recurrent neural network architecture. The RNN can be designed to capture the temporal and spatial dynamics of the neural populations by incorporating feedback loops that allow for the modeling of recurrent interactions among excitatory and inhibitory neurons. The model parameters, such as synaptic weights, time constants and effective connectivity weights, can be initialized based on prior empirical findings or estimated from the data. We use a loss function that quantifies the difference between the predicted ERP waveforms generated by the RNN and the observed ERP data. This can be achieved through mean squared error (MSE) or other appropriate metrics that reflect the accuracy of the model’s predictions. The training process involves optimizing the model parameters using backpropagation through time (BPTT) [68, 69, 70], a technique well-suited for RNNs that accounts for the temporal dependencies in the data. For parameter estimation, we applied Whole-Brain Modeling in PyTorch (WhoBPyt), a machine learning-based approach, to fit individual brain network models. The same framework was employed in Momi et al. (2023) for task-based MEG data [67].

The integration of the Jansen-Rit model with RNN deep learning techniques provides valuable insights into the dynamics of neural populations and their interactions, particularly in the context of ERPs such as the N400. Here, we outline three key insights derived from this modeling approach, focusing on local gains for pyramidal-excitatory-inhibitory (PEI) interactions, network dynamics represented through graph theory, and state responses from pyramidal, excitatory, and inhibitory populations.

The JR model allows for the examination of local gains within the neural populations, specifically focusing on the interactions between pyramidal (P), excitatory (E), and inhibitory (I) neurons. These local gains reflect how changes in synaptic efficacy among these populations can influence the overall neural dynamics. For instance, increased excitatory input to pyramidal cells can enhance their firing rates, leading to a more pronounced N400 response during semantic processing. Conversely, heightened inhibitory activity can dampen the response, indicating a potential imbalance in E-I dynamics. This local perspective is crucial for understanding how specific neural interactions contribute to the generation of ERPs and can inform the identification of biomarkers for psychosis risk.

The incorporation of the Schaefer atlas [71, 72] facilitates the application of graph theory to analyze network dynamics among the lateral forward and backward connections. By examining how these network dynamics influence the N400 response, researchers can gain insights into the broader organizational principles of brain function and how disruptions in network connectivity may correlate with semantic processing deficits observed in CHRP individuals. For example, altered connectivity within the default mode network or fronto-parietal network may be linked to impaired contextual processing, which is crucial for effective semantic memory activation.

The neural state response from pyramidal (P), excitatory (E), and inhibitory (I) populations can be modeled to reflect both local and network dynamics, providing a comprehensive view of how these interactions manifest in observable ERPs. By simulating the collective activity of these populations across the 200 regions defined by the Schaefer atlas, researchers can assess how local excitatory and inhibitory influences interact with the broader network context to generate the N400 waveform. This joint perspective is particularly relevant for understanding how reduced N400 amplitudes in CHRP individuals may indicate a failure to integrate contextual information effectively, leading to poorer social functioning outcomes. The model can also help identify specific regions or networks where E-I imbalances are most pronounced, offering potential targets for treatment intervention.

The following **local gain parameters** can be estimated from this model:

- **Pyramidal Excitation (Parameter A /** *H*_*e*_) – Represents the strength of excitatory input to pyramidal neurons. CHRP individuals demonstrated increased *A* values, reflecting heightened pyramidal excitability, consistent with cortical disinhibition.
- **Inhibitory Gain (Parameter B /** *H*_*i*_) – Captures the amplification of inhibitory interneuron activity. CHRP participants exhibited reduced inhibitory gain, suggesting weaker interneuron-mediated regulation of pyramidal output, which may contribute to inefficient semantic processing.
- **Pyramidal-to-Inhibitory Connectivity (Parameter C3)** – Reflects the influence of pyramidal neurons on inhibitory interneurons. A key finding was reduced *C*_3_ values in CHRP individuals, indicating a breakdown in feedback inhibition, which correlated with worsening social and role functioning over one year.
- **Excitatory-to-Pyramidal Connectivity (Parameter C2)** – Represents excitatory input from spiny stellate cells to pyramidal neurons. Dysregulation of *C*_2_ may impact how CHRP individuals process contextual predictions in semantic memory.
- **Pyramidal-to-Excitatory Connectivity (Parameter C1)** – Governs excitatory interactions among pyramidal neurons. Changes in *C*_1_ values may contribute to altered recurrent excitation, a feature linked to abnormal predictive coding in psychosis.
- **Decay Rate of Excitatory Postsynaptic Potential (Parameter** *a* **or** 1*/τ*_*e*_) – Determines how quickly excitatory potentials dissipate over time. Slower decay rates in CHRP individuals could indicate prolonged excitatory activity, affecting the integration of semantic information.
- **Decay Rate of Inhibitory Postsynaptic Potential (Parameter** *b* **or** 1*/τ*_*i*_) – Governs the temporal dynamics of inhibition. A reduction in *b* suggests delayed inhibitory processing, further exacerbating E-I imbalance.

### 2.5. Jansen–Rit Neural Mass Model and PCA of Inferred Neural States

In order to interpret the complex dynamics of cortical activity, we use a two-step approach. First, the Jansen–Rit (JR) neural mass model is employed to simulate the state responses in individual cortical regions. Second, Principal Component Analysis (PCA) is applied to the inferred neural states to extract the principal modes of activity across subjects and experimental conditions.

#### 2.5.1. Mathematical Formulation of the Jansen–Rit Model

The Jansen–Rit model is a well-established neural mass model that describes the dynamics of a cortical column via three interconnected neuronal populations: pyramidal neurons, excitatory interneurons, and inhibitory interneurons. Each population’s postsynaptic response to incoming action potentials is modeled by a second-order differential equation that is equivalent to a convolution with a synaptic impulse response function.

For a given neural population, the postsynaptic membrane potential *v*(*t*) in response to a presynaptic input *m*(*t*) is described by:

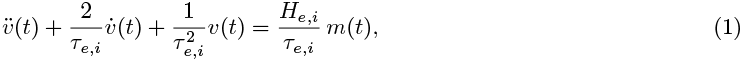

which is equivalent to the convolution

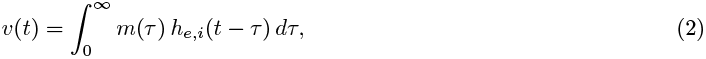

with the synaptic impulse response function (or pulse-to-wave operator) given by:

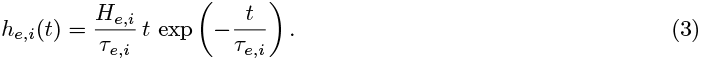

Here, *H*_*e,i*_ is the maximum postsynaptic potential and *τ*_*e,i*_ represents the effective time constant (with separate values for excitatory and inhibitory populations).

In addition, each population converts its membrane potential into an average firing rate via a sigmoidal (wave-to-pulse) operator:

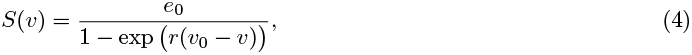

where *e*_0_ is the maximum firing rate, *r* determines the steepness of the sigmoid, and *v*_0_ is the potential at which half the maximum firing rate is achieved.

To facilitate numerical simulation, the second-order differential equations are reformulated as a set of first-order equations. For each cortical area *j* (with *j* = 1, …, *N*, where *N* = 200 in our whole-brain model), the JR model is expressed as:

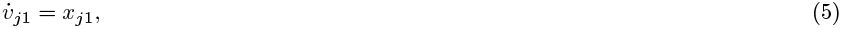

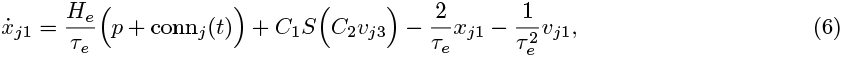

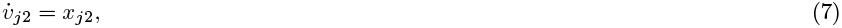

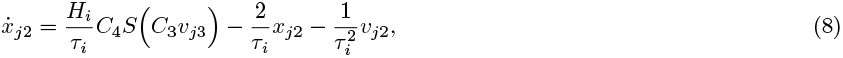

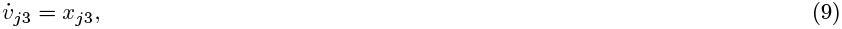

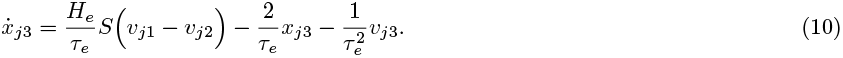

In these equations, *v*_*j*1_, *v*_*j*2_, and *v*_*j*3_ represent the average postsynaptic potentials of the excitatory interneurons, inhibitory interneurons, and pyramidal cells, respectively. The constant *p* represents an external perturbation—such as that induced by TMS—applied to the excitatory interneuron population. The term conn_*j*_(*t*) accounts for the excitatory input arriving at area *j* from all other areas in the network:

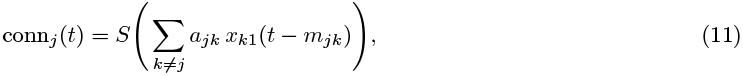

where *a*_*jk*_ is the connectivity weight from area *k* to area *j* and *m*_*jk*_ represents the conduction delay associated with that connection.

### 2.6. Integration with PCA for Dimensionality Reduction

The neural states generated by the JR model across the 200 cortical regions and under various experimental conditions result in high-dimensional data. To extract meaningful patterns, PCA is applied to these inferred state responses. The process involves:

1. **Concatenation and Normalization**: For each subject and condition, the normalized state responses—such as *P, E, I, P*_*v*_, *E*_*v*_, and *I*_*v*_—are concatenated into a signal vector:

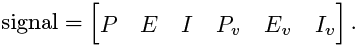

Each state is normalized as follows:

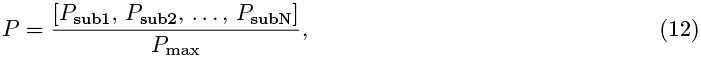

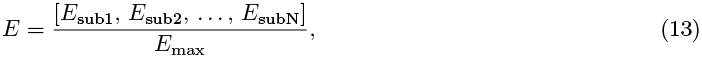

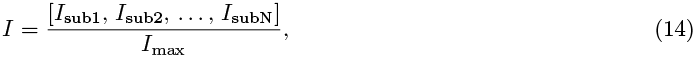

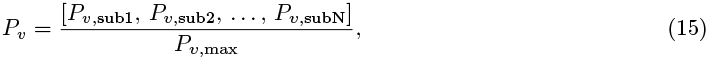

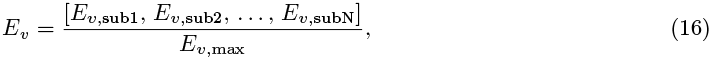

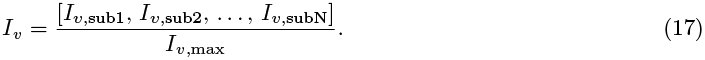
2. **Covariance Computation**: The covariance matrix of the aggregated signal vectors is computed as:

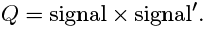
3. **Singular Value Decomposition (SVD)**: SVD is applied to the covariance matrix:

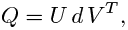

where the first column of *U*, denoted *u*_1_, corresponds to the direction of maximum variance.
4. **Projection**: Each state time series is projected onto the principal component space:

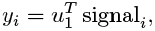

thereby reducing the dimensionality of the data while retaining the most informative features.

Maintaining the consistency of the eigenvectors across conditions is critical. This consistency ensures that the principal components reliably capture the dominant modes of neural dynamics, facilitating robust comparisons across experimental groups and conditions. Moreover, variations in the principal components may be correlated with behavioral or clinical outcomes, suggesting that these components can serve as biomarkers for underlying neurophysiological states.

In summary, the integration of the Jansen–Rit neural mass model with PCA provides a powerful framework for simulating and analyzing cortical dynamics. The JR model mathematically encapsulates the interplay between excitatory and inhibitory populations within cortical columns, while PCA reduces the resulting high-dimensional state responses into a set of dominant modes. This combined approach not only enhances the interpretability of complex neural data but also paves the way for identifying potential biomarkers in clinical applications, such as in the study of psychosis risk.

### 2.7. Partial Least Squares Analysis

Mean-centered task or behavioural partial least squares correlation (PLSC) analysis was used to capture model parameters that maximally represented differences between conditions or that maximally correlated with role and social functioning at baseline and 1-year later [73]. Singular value decomposition (SVD) was applied to the mean-centered matrix of parameter estimates. SVD re-expresses this matrix as a set of orthogonal singular vectors or latent variables (LVs), the number of which is equivalent to the total number of conditions. The LVs can be understood analogous to principal components in principal component analsys and account for the covariance of the original mean-centered matrix in decreasing order of magnitude. Statistical significance of LVs and reliability of region loadings on the LVs were assessed using permutation tests with 2000 permutations and bootstrapping with 2000 samples, respectively.

## 3. Results

### 3.1. Model Identifiability

To examine and ensure parameter identifiability and recovery, we combined three complementary safeguards during inversion. First, we performed *repeated inversions* (30 independent optimisations per ERP) and averaged the inferred parameters to reduce sensitivity to local minima and stochastic variation in the parameter landscape. Second, we optimised a *variational free energy* (ELBO) objective rather than mean-squared error alone, thus penalising model complexity and enforcing consistency with priors. Third, we imposed *weakly informative priors* on learnable parameters to confine solutions to biologically plausible regimes (e.g., gains, synaptic decay constants), thereby enhancing numerical stability and physiological interpretability. Together, these steps helped regularise estimation and approximate the posterior mean under the model.

### 3.2. Evidence from prior-to-posterior contraction

We quantified identifiability by examining prior and posterior marginals for each parameter and group. Across parameters, posterior distributions were systematically narrower than the priors, indicating substantial information gain from the ERP data and strong data-driven constraints on parameter values. On average, posterior standard deviations were approximately half of the prior (prior *σ* ≈ 1.0, posterior *σ* ≈ 0.5), consistent with appreciable contraction of uncertainty. Notably, pyramidal gain *A*, inhibitory feedback *C*_3_, and inhibitory decay *b* exhibited pronounced posterior sharpening, with separable posterior locations for CHRP and controls. These findings indicate that the data constrain parameters beyond prior expectations and support parameter-level identifiability.

To test whether credible intervals reflected unique solutions rather than compensatory trade-offs, we computed posterior correlation matrices separately for controls and CHRP using a common colour scale. Off-diagonal dependencies were uniformly small in both groups: no element exceeded |*r*| = 0.08, and the mean absolute correlation was *<* 0.03. The near-diagonal structure indicates minimal multicollinearity and successful disentanglement of parameters such as *A, b*, and *C*_3_, reinforcing interpretability of individual circuit mechanisms.

Beyond contraction, several parameters displayed *group-shifted* posteriors (e.g., higher *A* and faster *b* in CHRP), consistent with hypothesised cortical disinhibition and lending face validity to the mechanistic readouts. Because posterior covariances were negligible, these between-group shifts cannot be trivially attributed to parameter trade-offs, strengthening the case for biologically meaningful differences in local gain control and feedback inhibition.

In conclusion, (i) ensemble inversion under an ELBO objective with physiologically grounded priors, (ii) robust prior-to-posterior contraction, and (iii) uniformly low posterior inter-parameter correlations jointly demonstrate high practical identifiability of the JR model in this dataset. This supports the use of parameter estimates-particularly *A, b*, and *C*_3_-as reliable markers of altered excitation-inhibition balance in CHRP.

### 3.3. Model Face Validity

The neural states extracted from the model reflect the inferred neural activity at the population level. We specifically focused on the E/I balance, or the absolute value of the difference between the excitatory and inhibitory neural populations, and identified several peaks at 150 ms (peak 1), an undershooting response at 250 ms (peak 2), and a second positive peak at 350-450 ms (peak 3).

To evaluate the reliability and robustness of the Jansen-Rit neural mass model in characterizing N400 ERP responses, we conducted a behavior PLS analysis of the model parameters and the N400 ERP responses across healthy controls and CHRP groups. By resampling the data with replacement, bootstrap procedures estimate confidence intervals and identify parameter sets that remain stable despite sampling variability.

Figure 5 depicts the bootstrap ratio weighted (BSW)-results comparing semantic relatedness conditions (unrelated vs. related) in the long stimulus-onset asynchrony (SOA) condition. The upper panels show a significant contrast (p = 0.027) for HC participants, where several parameters, including inhibitory interneuron gains (*Iv*) and excitatory populations (*E*), exceed the critical reliability threshold (*±*2.5, indicated by red dashed lines). Parameters surpassing this threshold demonstrate high stability and serve as robust predictors of differences in the underlying neural dynamics. In contrast, the lower panels including the effects in CHRP individuals (p = 0.22) do not exhibit any parameters above the threshold, confirming the specificity and reliability of the observed effects in the significant condition.

**Figure 5:**
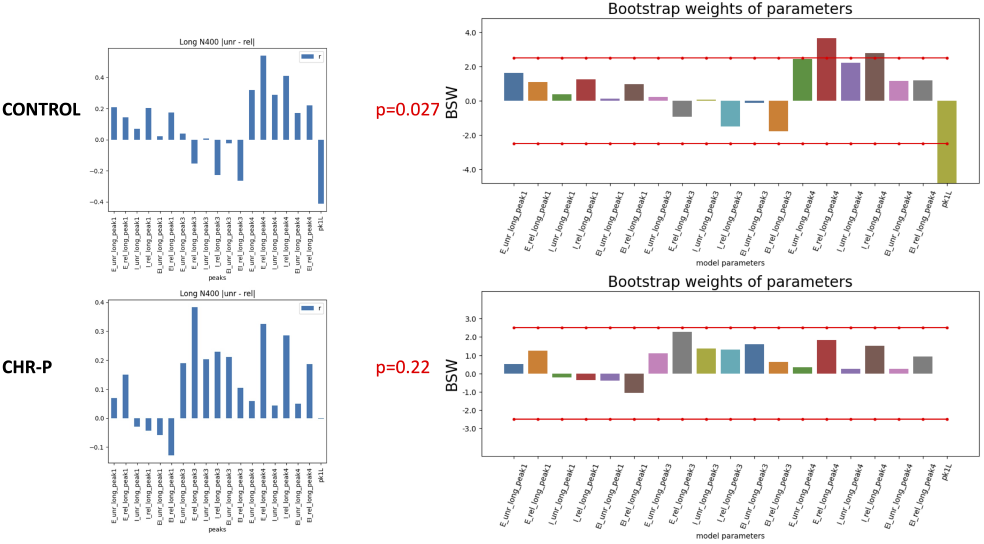
Model Face Validity: Behavior PLS analysis of the Jansen-Rit model parameters and N400 ERPs comparing semantic relatedness conditions (unrelated vs. related) under a long stimulus-onset asynchrony (SOA). **Top panels (p = 0.027):** Several parameters, including inhibitory interneuron gains (*Iv*) and excitatory populations (*E*), exceed the critical threshold (± 2.0, red dashed lines), indicating robust and reliable differentiation between unrelated and related conditions. **Bottom panels (p = 0.22):** No parameters surpass the threshold in the CHRP group, confirming the specificity of the observed effects in the control group. These findings highlight the stability of the implicated parameters and underscore the model’s reliability for probing neurocomputational mechanisms underlying semantic processing.

To further examine the model’s performance, we performed the same BSW analysis for the **short** SOA condition. In this case, neither the control group (p = 0.37) nor the clinical high-risk (CHRP) group (p = 0.08) showed correlations between the neural state parameters and N400 ERPs that exceeded the *±*2.5 threshold. This indicates that, at the short SOA, there are no robust or highly stable differences in the neural mass model parameters related to semantic processing for either group. The absence of parameters exceeding the *±*2.5 bootstrap-weight threshold in the short SOA condition suggests that semantic processing in this temporal regime is not strongly differentiated by the neurocomputational mechanisms captured in the Jansen–Rit model for either controls or CHRP participants. Short SOAs (≈ 250–300 ms) primarily index automatic spreading activation in semantic networks, a process that is relatively resistant to higher-order contextual influences and is thought to be less impaired in psychosis-spectrum populations. Because automatic priming operates rapidly and with minimal reliance on controlled integration or working-memory maintenance, the model’s local gain parameters—particularly those reflecting excitatory–inhibitory (E–I) balance and synaptic time constants—may contribute less variance to the N400 signal in this time window.

These findings underscore the model’s sensitivity to key neural mass parameters involved in semantic processing. The stability of these parameter estimates across bootstrap resamples substantiates the model’s validity and enhances confidence that differences in excitatory and inhibitory population dynamics are not artifacts of particular samples. Consequently, the bootstrap-weight analysis reinforces the suitability of the Jansen-Rit model for investigating neurocomputational mechanisms underlying cognitive processes, including those relevant to clinical high-risk populations.

### 3.4. Key Temporal Peaks Analysis

The early 150 ms peak is often associated with initial sensory processing and the engagement of attentional mechanisms. It reflects the brain’s rapid response to semantic stimuli and may indicate the efficiency of early cognitive processing. Second, the 250 ms peak is particularly noteworthy as it represents an undershooting response, which may indicate a failure to adequately predict or integrate semantic information. This undershooting can be indicative of cognitive disruptions, particularly in CHRP populations. Third, the 350 ms and 450 ms peaks are typically associated with the N400 component, reflecting the brain’s processing of semantic incongruities. The amplitude of the N400 is sensitive to the semantic relationship between prime and target stimuli, and reductions in this amplitude have been linked to deficits in semantic processing in CHRP individuals. Finally, the later peak at 750 ms is associated with integration of semantic information and may reflect the cognitive load associated with processing complex stimuli.

### 3.5. Main Effects of Condition

Analysis of stimulus onset asynchrony (SOA) effects on local gain parameters using Partial Least Squares (PLS) analysis revealed significant differences in excitatory and inhibitory dynamics across short (300 ms) and long (750 ms) SOA trials (Figure 6). PLS analysis demonstrated a robust main effect of SOA, with long SOA conditions (REL-Long, UNR-Long) exhibiting larger excitatory gain (A) and inhibitory decay rate (b), whereas short SOA conditions (REL-Short, UNR-Short) showed greater pyramidal-to-inhibitory connectivity (C4) and inhibitory gain (B) (p = 0.001). Variable Importance in Projection (VIP) scores confirmed A, b, C4, and B parameters as the strongest predictors of SOA-related differences, emphasizing the interplay between excitatory and inhibitory mechanisms in adapting to temporal constraints on semantic processing. These findings suggest that long SOA trials facilitate predictive semantic integration through increased excitatory and decay dynamics, whereas short SOA trials rely more on inhibitory control mechanisms to regulate early-stage automatic processing.

**Figure 6:**
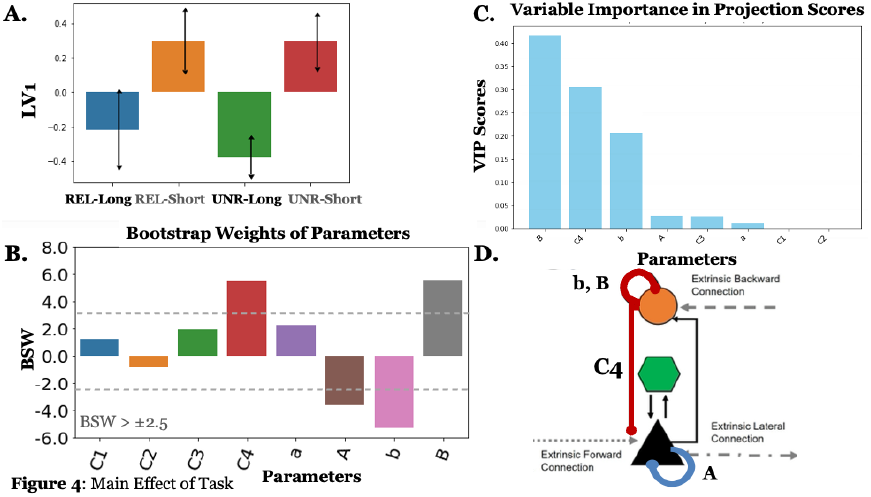
Effect of Stimulus Onset Asynchrony (SOA) on Local Gain Parameters: Partial Least Squares (PLS) analysis revealed significant differences in excitatory and inhibitory dynamics between short (300 ms) and long (750 ms) SOA conditions. (A) The first LV shows a main effect of SOA, with larger excitatory gain (A) and inhibitory decay rate (b) in long SOA trials (REL-Long, UNR-Long), whereas short SOA trials (REL-Short, UNR-Short) exhibited greater pyramidal-to-inhibitory connectivity (C4) and inhibitory gain (B). (B) Bootstrap weights over model parameters show reliable SOA-related differences, with C4 and B dominating in short SOA conditions, while A and b are more pronounced in long SOA conditions. (C) Variable Importance in Projection (VIP) scores confirm that A, b, C4, and B are the strongest predictors of SOA-related effects. (D) Schematic representation of the neural mass model highlights the role of excitatory gain (A) and inhibitory decay rate (b) in long SOA trials, supporting predictive semantic integration, while short SOA trials rely on increased inhibitory regulation (C4, B) to constrain automatic activation. These findings suggest that SOA critically modulates excitation-inhibition (E-I) balance during semantic processing, with short SOA trials emphasizing rapid inhibitory control and long SOA trials favoring excitatory mechanisms for sustained integration.

### 3.6. Group Differences in Local Gain Parameters

Neurocomputational modeling of N400 ERPs using the Jansen-Rit (JR) model revealed significant group differences in local gain parameters between CHRP individuals and healthy controls, highlighting disruptions in excitatory-inhibitory (E-I) balance consistent with NMDAR hypofunction in emerging psychosis. CHRP individuals exhibited heightened excitatory drive in pyramidal cells (Parameter A; Cohen’s d=0.59, p=0.01) across all experimental conditions, suggesting increased stimulus-driven excitation relative to controls (see Figure 7). This aligns with theories of cortical disinhibition, where reduced NMDA-mediated inhibitory control results in excessive pyramidal firing.

**Figure 7:**
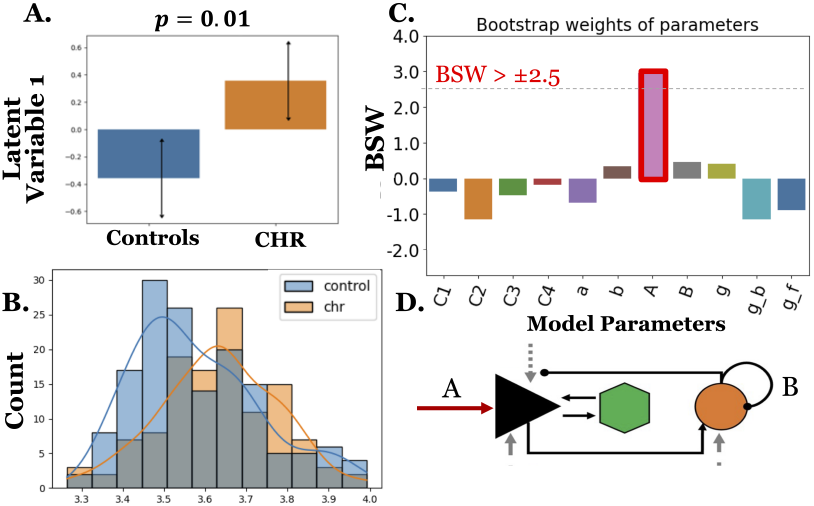
Main Effect of Group on Local Gain Parameters: Neurocomputational modeling of N400 ERP data using the Jansen-Rit (JR) model showed greater excitation of pyramidal cells in response to target word stimuli across all conditions (prime-target SOA and relatedness) in CHRP patients (*n* = 46) compared to controls (*N* = 38). (**A**) Latent variable 1 (from partial least squares analysis) reveals a significant group difference. (**B**) LV1 score distribution for controls and CHRP across different model initializations highlights a consistent shift in CHRP scores relative to controls. (**C**) Bootstrap weights identify parameter A (BSW *> ±*2.5, red box) as reliable. (**D**) Schematic representation of the neural mass model.

### 3.7. Time-resolved excitation–inhibition (E–I) trajectories

Principal-component projection of the absolute difference between excitatory and inhibitory states (|*E*−*I*|) uncovered four reproducible peaks: early sensory (150ms), undershoot (250ms), classical N400 (350–450ms), and late integration (750ms). Long-SOA, unrelated trials (peak1, 70–100ms). CHRP participants displayed a sharply elevated |E–I| burst relative to controls (LV1, p=0.0004), driven by increased excitability - i.e., increased parameter A and strengthened excitatory-to-pyramidal connectivity (C2), and accelerated inhibitory decay, collectively indicating early cortical disinhibition.

Short-SOA, related trials (peak 3, 350ms). Conversely, CHRP exhibited a larger N400-window |E–I| peak sustained by enhanced inhibitory-to-pyramidal feedback (C3) and prolonged inhibitory decay (b) (LV1, p=0.003). This pattern reflects a temporally specific inhibitory reinforcement that counters local hyper-excitation.

Late integration peak (750ms). Both groups exhibited a ∼ 750-ms rise in |*E* − *I*|, but the amplitude was 18% lower in CHRP (*t* = 2.4, *p* = 0.018), potentially suggesting inefficient semantic consolidation.

### 3.8. Relationship with Psychosocial Functioning

In the long-SOA, unrelated condition, the estimated model-based excitation–inhibition magnitude (E-I) showed a sharply elevated early peak (70-100 ms) in CHRP relative to controls. Multivariate partial-least-squares confirmed a robust group separation (LV1,p=0.0004), with increased excitation relative to inhibition in CHRP compared to controls. These effects are driven by increased pyramidal EPSP gain (parameter A), strengthened excitatory-to-pyramidal connectivity, and faster inhibitory decay—together indicating local cortical disinhibition. Critically, greater amplitude of this early (E–I) peak predicted worse social functioning at one-year follow-up (*r* = −.56, *p* = .003; Figure 8 lower-right). Conversely, when prime-to-target SOA was 300ms and the words were semantically related, the magnitude of the model-derived excitation–inhibition signal (E – I) showed a larger N400 time-window peak (350ms; peak3) in CHRP relative to controls. PLS analysis confirmed a reliable group separation (LV1, p=0.003; Figure 9 upper-centre) that was driven by two parameter clusters: (i) stronger inhibitory-to-pyramidal feedback (C3) and (ii) lengthened inhibitory post-synaptic decay (parameter b). Together, these features point to a slowing yet strengthening of inhibitory control that counteracts local hyper-excitation in the prodrome (*r* = .60, *p* = .001), suggesting that this inhibitory reinforcement may confer functional resilience despite emerging risk.

**Figure 8:**
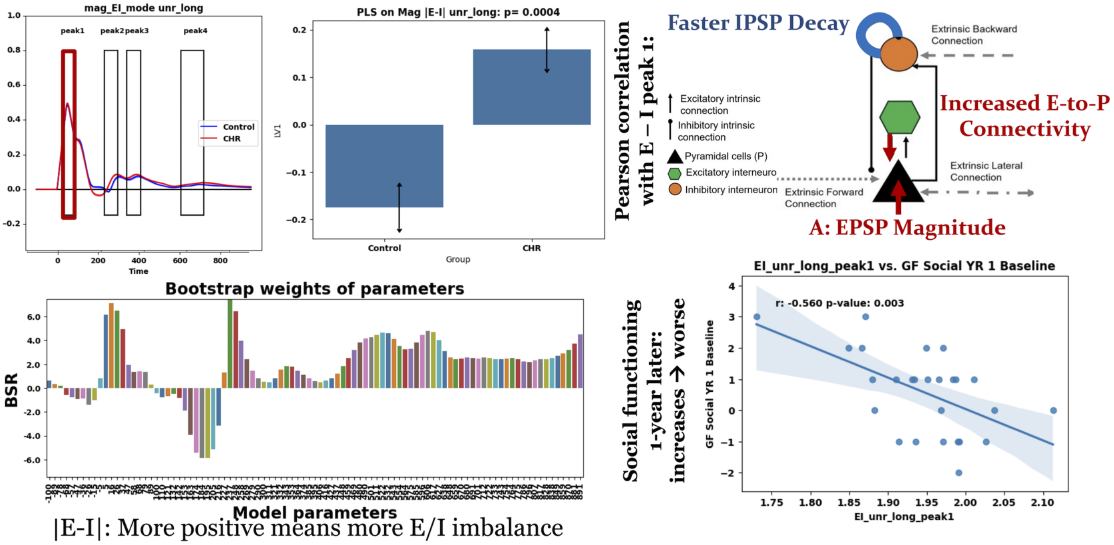
Early sensory disinhibition predicts poorer social functioning in CHRP. (*Top-left*) Time–course of the absolute excitation–inhibition difference (|*E − I*|) during the long-SOA/unrelated condition. The first peak (70–100ms; red box) is markedly larger in CHRP (red) than in controls (blue). (*Top-centre*) Latent-variable scores from a partial least squares analysis confirm a significant main effect of group on this early |*E − I*| magnitude (*p* = 0.0004). (*Top-right*) Circuit schematic illustrating the parameter constellation driving the disinhibitory burst: faster IPSP decay, larger pyramidal EPSP magnitude (*A*) and increased excitatory-to-pyramidal coupling. (*Bottom-left*) Bootstrap ratios of model parameters; positive bars highlight gains that increase the |*E − I*| imbalance. (*Bottom-right*) Early disinhibitory peak amplitude negatively correlates with Global Functioning–Social (GFS) scores one year later (*r* = −0.56, *p* = 0.003), indicating that stronger early cortical disinhibition forecasts poorer social outcome.

**Figure 9:**
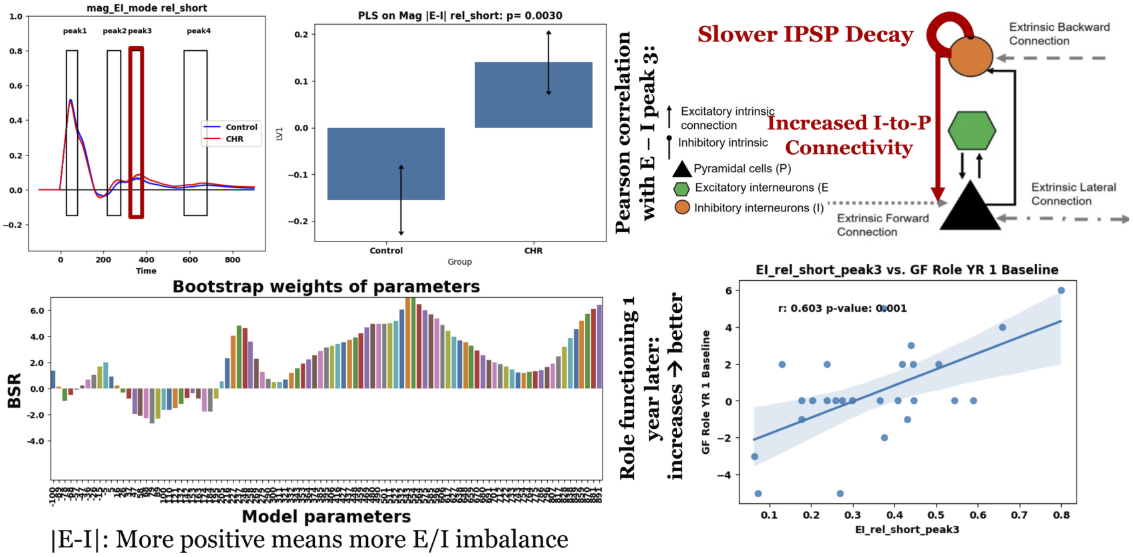
Later N400-window inhibitory reinforcement predicts better role functioning in CHR-P. (*Top-left*) |*E − I*|trajectories for the short-SOA/related condition. The third peak (350ms; red box) is accentuated in CHR-P relative to controls. (*Top-centre*) Partial least squares scores demonstrate a robust group effect on this later inhibitory peak (*p* = 0.003). (*Top-right*) Schematic highlights the underlying mechanism: slower IPSP decay and stronger inhibitory-to-pyramidal feedback enhance late inhibition. (*Bottom-left*) Bootstrap ratios reveal parameter clusters that elevate (positive bars) or dampen (negative bars) the late |*E − I*| response. (*Bottom-right*) Larger late inhibitory peaks correlate positively with Global Functioning–Role (GFR) scores at one-year follow-up (*r* = 0.60, *p* = 0.001), suggesting that the capacity to recruit delayed inhibition supports better occupational functioning.

## 4. Discussion

This study integrates high-density EEG, connectome-constrained neural-mass modelling, and prospective functional outcome data to elucidate the circuit mechanisms underlying N400 abnormalities in youth at clinical high risk for psychosis (CHRP). Two principal insights emerge.

### 1. Pervasive excitatory dominance with temporally specific signatures

Across all conditions, CHRP participants displayed heightened pyramidal excitatory gain and reduced inhibitory gain, resulting in a global shift toward cortical disinhibition. Yet this imbalance was not uniform across time: an early sensory burst of excitation (70–100ms) dominated during semantic processing, whereas a later, context-dependent disinhibition emerged in the N400 window (350ms). These findings demonstrate that psychosis-risk physiology is best characterised by dynamic, not static, perturbations in excitation–inhibition E-I balance.

### 2. Neurocomputational biomarkers predict functional trajectories

The amplitude of the early excitatory peak prospectively predicts poorer social functioning, whereas the magnitude of the later inhibitory peak predicted better role functioning one year later. Together with local-gain parameters, these indices explained more than half of the variance in psychosocial outcome, substantially exceeding traditional ERP measures.

By linking aberrant semantic integration to precise circuit motifs, this work bridges cognitive electrophysiology with neurocomputational mechanisms reinforcing the centrality of E–I balance in psychosis pathophysiology [74, 37]. This micro-circuit profile in consistent with the broader ERP literature in which N400 attenuation is already detectable in the prodromal phase and deepens after transition to psychosis [29, 30, 32, 75].

Previous studies have shown that individuals at clinical high risk for psychosis (CHRP) reliably demonstrate reduced N400 semantic-priming effects relative to healthy controls and typically occupy an intermediate position between controls and patients with first-episode psychosis (FEP) or chronic schizophrenia spectrum disorders (SSD)-consistent with a spectrum-wide gradient: healthy *>* CHRP *>* FEP ≈ SSD [76, 16, 24, 77]. Additionally, timing constraints matter: N400 abnormalities in CHRP and the schizophrenia spectrum are most robust under longer prime–target intervals, with relatively preserved automatic priming at brief SOAs, implicating deficits in controlled semantic integration rather than initial activation of semantic processing - precisely the SOA dissociation we and others have observed [32, 78, 75].

It is important to note, however, that long SOA deficits in CHRP are still modest compared to those with frank psychosis. SSD patients show distinctive N400 abnormalities depending on the SOA. At long SOAs (≥ 500 ms), patients consistently show reduced priming effects – their N400 response to related targets is more negative, resulting in a blunted or even absent N400 difference between related and unrelated words [24]. By contrast, at short SOAs (≈250–300 ms) previous studies have found that SSD patients’ N400 priming is relatively preserved. In fact, at short SOAs, SSD patients often show greater than normal N400 reduction for related words, which has been attributed to an unrestrained activation in semantic memory [19, 21, 22]. These SOA-dependent patterns support the idea that psychosis patients fail to appropriately generate or apply top-down contextual predictions [78, 32, 24, 27]. Our finding that deficits are most evident at longer SOAs thus replicates earlier behavioural-EEG work demonstrating that CHRP individuals struggle primarily with controlled contextual integration, whereas more automatic priming is relatively spared.

Beyond group differences, CHRP studies have examined correlations between N400 measures and clinical outcomes even before psychosis onset [30, 32, 25]. Notably, N400 deficits in CHRP correlate with concurrent cognitive and functional impairments [78, 30]. In one sample of 35 CHR patients, smaller N400 priming at 300ms SOA was linked to poorer role (academic/work) functioning, and a reduced N400 priming effect at 750ms was associated with lower global cognitive performance (MATRICS battery scores) [78].

#### 4.0.1. Predictive coding and NMDA-hypofunction mechanisms

Interpreting the N400 as a lexico-semantic prediction-error signal situates our excitation–inhibition findings squarely within predictive-coding accounts of psychosis, in which aberrant precision weighting weakens top-down contextual constraints, yielding reduced N400 amplitudes [25] and diminished belief updating [79]. SOA-dependent N400 abnormalities across CHRP, FEP, and schizophrenia strongly support this framework. In a healthy brain, context derived from prior input and world knowledge shapes predictions about upcoming words, allowing expected items to elicit attenuated N400 responses. In psychosis-spectrum conditions, however, long-SOA paradigms reveal a consistent inability to suppress N400 for related words: even predictable inputs register as surprising, reflecting a failure to use priors to minimise prediction error [23, 21]. This pattern echoes classic context-integration theories, in which deficits in maintaining contextual information in working memory and integrating it with new input undermine controlled semantic processing [80, 81]. Our model-derived profile of heightened pyramidal gain coupled with delayed feedback inhibition is consistent with NMDA-receptor hypofunction on parvalbumin interneurons, a leading molecular model of prodromal pathophysiology capable of reproducing N400 priming deficits in silico [82, 83]. Such a mechanism could explain why context-driven prediction is weak or noisy from the prodromal stage onward, yielding the “cloudy semantic crystal ball” that manifests clinically as subtle speech anomalies or idiosyncratic interpretations in CHRP, and as incoherent thought or delusions in established illness [19, 30, 78]. Occasional hyperpriming at short SOA fits within this account as a failure to appropriately constrain priors: overly diffuse, weak-precision predictions permit broad automatic activation but fail to focus expectations during controlled integration [84, 85]. Overall, the convergence of SOA-manipulation findings, predictive-coding theory, and our circuit-level results strengthens the view that reduced use of context and aberrant prediction error signalling are core mechanistically grounded features of psychosis from its earliest phase [86, 87].

#### 4.0.2. Clinical implications

Cross-sectionally, attenuated N400 reliably differentiates CHRP from healthy controls, underscoring its potential as a state biomarker of emerging psychosis-related pathology [75, 29]. Our neurocomputational modelling results extend these ERP findings by linking SOA-dependent N400 differences to specific excitation–inhibition mechanisms [37]. In long SOA with targets unrelated to primes, CHRP participants exhibited an early sensory disinhibition (70–100 ms) driven by increased pyramidal excitatory gain, strengthened excitatory-to-pyramidal connectivity, and faster inhibitory decay, consistent with NMDA-receptor hypofunction and cortical disinhibition in the psychosis prodrome. Crucially, the amplitude of this early E-I burst predicted poorer social functioning at one-year follow-up (*r* = −0.56), demonstrating mechanistic and prognostic relevance. In contrast, during short-SOA, related trials, CHR-P showed a larger N400-window inhibitory peak (∼350 ms) sustained by stronger inhibitory-to-pyramidal feedback and prolonged inhibitory decay, which correlated positively with role functioning (*r* = 0.60), suggesting that temporally specific inhibitory reinforcement may confer functional resilience. At the late integration stage (∼750 ms), CHR-P displayed reduced E-I amplitude relative to controls, potentially reflecting inefficient semantic consolidation. Here, semantic consolidation denotes the within-trial, late-stage stabilization and strengthening of an integrated, context-appropriate meaning representation, operationalized in our model as the ∼750 ms settling of network dynamics that binds prime–target semantics and supports subsequent retrieval.

These findings provide a mechanistic account of the SOA effects observed in conventional ERP analyses: long SOA facilitates predictive semantic integration through heightened excitatory and decay dynamics, which in CHRP is pathologically exaggerated early in processing; short SOA relies more on inhibitory regulation to constrain automatic activation, which CHRP may recruit adaptively in certain contexts. Longitudinal CHRP cohorts have yielded mixed results for predicting transition to psychosis, where baseline N400 measures do not reliably separate converters from non-converters over two years [32], but our modelling suggests that parameter-specific E–I signatures at defined temporal windows may hold greater short-term prognostic utility for functional outcomes.

Where the N400, and now its model-derived E–I correlates, show particular promise is in reflecting functional prognosis. Across CHRP and chronic schizophrenia, reduced N400 is associated with poorer real-world functioning [78], and our data further pinpoint circuit-level markers that track with both impairment and resilience. In clinical applications, the N400 is unlikely to function as a diagnostic “silver bullet,” but integrated into multimodal risk-stratification frameworks alongside other EEG measures (e.g., mismatch negativity) [88, 89, 90] and neuroimaging measures [91], it could help identify individuals who might benefit from targeted early interventions. Moreover, longitudinal tracking of both ERP- and model-derived measures could inform treatment monitoring where normalization of early disinhibitory bursts or enhancement of compensatory inhibitory peaks may index a positive shift in predictive processing.

Our study demonstrates that dynamic E–I disturbances, which characterised by early cortical disinhibition and context-dependent compensatory inhibition, underlie N400 impairments in CHRP and hold promise as prognostic indicators. The multimodal, computational framework presented here offers a scalable path to precision diagnostics and targeted therapeutics in the psychosis prodrome.

## Data Availability

All data produced in the present study are available upon reasonable request to the authors.

## Notes

### Competing Interest Statement

The authors have declared no competing interest.

### Funding Statement

This study was funded by the Canadian Institutes of Health Research.

### Author Declarations

Ethics committee/IRB of the Centre for Addiction and Mental Health affiliated with the University of Toronto gave ethical approval for this work.

## References

1. McGrath J, Saha S, Chant D, Welham J. Schizophrenia: A Concise Overview of Incidence, Prevalence, and Mortality. Epidemiologic Reviews 2008;30(1):67–76. URL: https://doi.org/10.1093/epirev/mxn001. doi:10.1093/epirev/mxn001.

2. van Os J, Kapur S. Schizophrenia. The Lancet 2009;374(9690):635–45. URL: http://www.sciencedirect.com/science/article/pii/S0140673609609958. doi:10.1016/S0140-6736(09)60995-8.

3. Goeree R, Farahati F, Burke N, Blackhouse G, O’Reilly D, Pyne J, Tarride JE. The economic burden of schizophrenia in Canada in 2004. Current Medical Research and Opinion 2005;21(12):2017–28. doi:10.1185/030079905X75087.

4. Addington J, Heinssen R. Prediction and prevention of psychosis in youth at clinical high risk. Annual Review of Clinical Psychology 2012;8:269–89. doi:10.1146/annurev-clinpsy-032511-143146.

5. Woods SW, Powers AR, Taylor JH, Davidson CA, Johannesen JK, Addington J, Perkins DO, Bearden CE, Cadenhead KS, Cannon TD, Cornblatt BA, Seidman LJ, Tsuang MT, Walker EF, McGlashan TH. Lack of Diagnostic Pluripotentiality in Patients at Clinical High Risk for Psychosis: Specificity of Comorbidity Persistence and Search for Pluripotential Subgroups. Schizophrenia Bulletin 2018;44(2):254–63. doi:10.1093/schbul/sbx138.

6. Beck K, Studerus E, Andreou C, Egloff L, Leanza L, Simon AE, Borgwardt S, Riecher-Rössler A. Clinical and functional ultra-long-term outcome of patients with a clinical high risk (CHR) for psychosis. European Psychiatry: The Journal of the Association of European Psychiatrists 2019;62:30–7. doi:10.1016/j.eurpsy.2019.08.005.

7. de Wit S, Schothorst PF, Oranje B, Ziermans TB, Durston S, Kahn RS. Adolescents at ultra-high risk for psychosis: long-term outcome of individuals who recover from their at-risk state. European Neuropsychopharmacology: The Journal of the European College of Neuropsychopharmacology 2014;24(6):865–73. doi:10.1016/j.euroneuro.2014.02.008.

8. Michel C, Ruhrmann S, Schimmelmann BG, Klosterkötter J, Schultze-Lutter F. Course of clinical highrisk states for psychosis beyond conversion. European Archives of Psychiatry and Clinical Neuroscience 2018;268(1):39–48. doi:10.1007/s00406-016-0764-8.

9. Polari A, Lavoie S, Yuen HP, Amminger P, Berger G, Chen E, deHaan L, Hartmann J, Markulev C, Melville F, Nieman D, Nordentoft M, Riecher-Rössler A, Smesny S, Stratford J, Verma S, Yung A, McGorry P, Nelson B. Clinical trajectories in the ultra-high risk for psychosis population. Schizophrenia Research 2018;197:550–6. doi:10.1016/j.schres.2018.01.022.

10. Carrión RE, McLaughlin D, Goldberg TE, Auther AM, Olsen RH, Olvet DM, Correll CU, Cornblatt BA. Prediction of functional outcome in individuals at clinical high risk for psychosis. JAMA psychiatry 2013;70(11):1133–42. doi:10.1001/jamapsychiatry.2013.1909.

11. Schmidt A, Cappucciati M, Radua J, Rutigliano G, Rocchetti M, Dell’Osso L, Politi P, Borgwardt S, Reilly T, Valmaggia L, McGuire P, Fusar-Poli P. Improving Prognostic Accuracy in Subjects at Clinical High Risk for Psychosis: Systematic Review of Predictive Models and Meta-analytical Sequential Testing Simulation. Schizophrenia Bulletin 2016;:sbw098URL: https://academic.oup.com/schizophreniabulletin/article-lookup/doi/10.1093/schbul/sbw098. doi:10.1093/schbul/sbw098.

12. Kim HS, Shin NY, Jang JH, Kim E, Shim G, Park HY, Hong KS, Kwon JS. Social cognition and neurocognition as predictors of conversion to psychosis in individuals at ultra-high risk. Schizophrenia Research 2011;130(1):170–5. URL: http://www.sciencedirect.com/science/article/pii/S0920996411002301. doi:10.1016/j.schres.2011.04.023.

13. Kim M, Lee TH, Yoon YB, Lee TY, Kwon JS. Predicting Remission in Subjects at Clinical High Risk for Psychosis Using Mismatch Negativity. Schizophrenia Bulletin 2018;44(3):575–83. doi:10.1093/schbul/sbx102.

14. Sawada K, Kanehara A, Sakakibara E, Eguchi S, Tada M, Satomura Y, Suga M, Koike S, Kasai K. Identifying neurocognitive markers for outcome prediction of global functioning in individuals with first-episode and ultra-high-risk for psychosis. Psychiatry and Clinical Neurosciences 2017;71(5):318–27. doi:10.1111/pcn.12522.

15. Bodatsch M, Brockhaus-Dumke A, Klosterkötter J, Ruhrmann S. Forecasting psychosis by event-related potentials-systematic review and specific meta-analysis. Biological Psychiatry 2015;77(11):951–8. doi:10.1016/j.biopsych.2014.09.025.

16. Kutas M, Federmeier KD. Thirty years and counting: Finding meaning in the N400 component of the event related brain potential (ERP). Annual review of psychology 2011;62:621–47. URL: https://www.ncbi.nlm.nih.gov/pmc/articles/PMC4052444/. doi:10.1146/annurev.psych.093008.131123.

17. Nobre AC, Allison T, McCarthy G. Word recognition in the human inferior temporal lobe. Nature 1994;372(6503):260–3. URL: https://www.nature.com/articles/372260a0. doi:10.1038/372260a0; number: 6503 Publisher: Nature Publishing Group.

18. Kutas M, Federmeier KD. Electrophysiology reveals semantic memory use in language comprehension. Trends in Cognitive Sciences 2000;4(12):463–70. doi:10.1016/s1364-6613(00)01560-6.

19. Condray R, Siegle GJ, Cohen JD, van Kammen DP, Steinhauer SR. Automatic activation of the semantic network in schizophrenia: evidence from event-related brain potentials. Biological Psychiatry 2003;54(11):1134–48. doi:10.1016/s0006-3223(03)00699-1.

20. Iakimova G, Passerieux C, Laurent JP, Hardy-Bayle MC. ERPs of metaphoric, literal, and incongruous semantic processing in schizophrenia. Psychophysiology 2005;42(4):380–90. doi:10.1111/j.1469-8986.2005.00303.x.

21. Kiang M, Kutas M, Light GA, Braff DL. An event-related brain potential study of direct and indirect semantic priming in schizophrenia. The American Journal of Psychiatry 2008;165(1):74–81. doi:10.1176/appi.ajp.2007.07050763; place: US Publisher: American Psychiatric Assn.

22. Kostova M, Passerieux C, Laurent JP, Saint-Georges C, Hardy-Baylé MC. Functional analysis of the deficit in semantic context processes in schizophrenic patients: an event-related potentials study. Neurophysiologie Clinique = Clinical Neurophysiology 2003;33(1):11–22. doi:10.1016/s0987-7053(03)00006-6.

23. Mathalon DH, Roach BJ, Ford JM. Automatic semantic priming abnormalities in schizophrenia. International Journal of Psychophysiology 2010;75(2):157–66. doi:10.1016/j.ijpsycho.2009.12.003; place: Netherlands Publisher: Elsevier Science.

24. Mohammad OM, DeLisi LE. N400 in schizophrenia patients. Current Opinion in Psychiatry 2013;26(2):196–207. doi:10.1097/YCO.0b013e32835d9e56.

25. Kiang M, Gerritsen CJ. The N400 event-related brain potential response: A window on deficits in predicting meaning in schizophrenia. International Journal of Psychophysiology 2019;145:65–9. doi:10.1016/j.ijpsycho.2019.04.005; place: Netherlands Publisher: Elsevier Science.

26. Besche-Richard C, Iakimova G, Hardy-Baylé MC, Passerieux C. Behavioral and brain measures (N400) of semantic priming in patients with schizophrenia: test-retest effect in a longitudinal study. Psychiatry and Clinical Neurosciences 2014;68(5):365–73. doi:10.1111/pcn.12137.

27. Kiang M, Prugh J, Kutas M. An event-related brain potential study of schizotypal personality and associative semantic processing. International journal of psychophysiology : official journal of the International Organization of Psychophysiology 2010;75(2):119–26. URL: https://www.ncbi.nlm.nih.gov/pmc/articles/PMC2827666/. doi:10.1016/j.ijpsycho.2009.10.005.

28. Kiang M, Kutas M. Association of schizotypy with semantic processing differences: An event-related brain potential study. Schizophrenia Research 2005;77(2-3):329–42. doi:10.1016/j.schres.2005.03.021; place: Netherlands Publisher: Elsevier Science.

29. Lepock JR, Mizrahi R, Korostil M, Maheandiran M, Gerritsen CJ, Drvaric L, Ahmed S, Bagby RM, Kiang M. N400 event-related brain potential evidence for semantic priming deficits in persons at clinical high risk for psychosis. Schizophrenia Research 2019;204:434–6. URL: https://www.sciencedirect.com/science/article/pii/S0920996418305462. doi:10.1016/j.schres.2018.08.033.

30. Lepock JR, Ahmed S, Mizrahi R, Gerritsen CJ, Maheandiran M, Drvaric L, Bagby RM, Korostil M, Light GA, Kiang M. Relationships between cognitive event-related brain potential measures in patients at clinical high risk for psychosis. Schizophrenia Research 2020;226:84–94. URL: https://www.sciencedirect.com/science/article/pii/S0920996419300143. doi:10.1016/j.schres.2019.01.014.

31. Kiang M, Kutas M, Light GA, Braff DL. Electrophysiological insights into conceptual disorganization in schizophrenia. Schizophrenia Research 2007;92(1-3):225–36. doi:10.1016/j.schres.2007.02.001.

32. Lepock JR, Sanches M, Ahmed S, Gerritsen CJ, Korostil M, Mizrahi R, Kiang M. N400 event-related brain potential index of semantic processing and two-year clinical outcomes in persons at high risk for psychosis: A longitudinal study. The European Journal of Neuroscience 2023;doi:10.1111/ejn.16074.

33. Bearden CE, Wu KN, Caplan R, Cannon TD. Thought disorder and communication deviance as predictors of outcome in youth at clinical high risk for psychosis. Journal of the American Academy of Child and Adolescent Psychiatry 2011;50(7):669–80. doi:10.1016/j.jaac.2011.03.021.

34. Spencer TJ, Thompson B, Oliver D, Diederen K, Demjaha A, Weinstein S, Morgan SE, Day F, Valmaggia L, Rutigliano G, De Micheli A, Mota NB, Fusar-Poli P, McGuire P. Lower speech connectedness linked to incidence of psychosis in people at clinical high risk. Schizophrenia Research 2021;228:493–501. doi:10.1016/j.schres.2020.09.002.

35. Sabb FW, van Erp TGM, Hardt ME, Dapretto M, Caplan R, Cannon TD, Bearden CE. Language network dysfunction as a predictor of outcome in youth at clinical high risk for psychosis. Schizophrenia Research 2010;116(2-3):173–83. doi:10.1016/j.schres.2009.09.042.

36. Grunwald T, Beck H, Lehnertz K, Blümcke I, Pezer N, Kurthen M, Fernández G, Van Roost D, Heinze HJ, Kutas M, Elger CE. Evidence relating human verbal memory to hippocampal N-methyl-D-aspartate receptors. Proceedings of the National Academy of Sciences of the United States of America 1999;96(21):12085–9. doi:10.1073/pnas.96.21.12085.

37. Krystal JH, Anticevic A, Yang GJ, Dragoi G, Driesen NR, Wang XJ, Murray JD. Impaired Tuning of Neural Ensembles and the Pathophysiology of Schizophrenia: A Translational and Computational Neuroscience Perspective. Biological Psychiatry 2017;81(10):874–85. URL: https://www.sciencedirect.com/science/article/pii/S0006322317300367. doi:10.1016/j.biopsych.2017.01.004.

38. Schmidt A, Diaconescu AO, Kometer M, Friston KJ, Stephan KE, Vollenweider FX. Modeling Ketamine Effects on Synaptic Plasticity During the Mismatch Negativity. Cerebral Cortex 2013;23:2394–406. doi:10.1093/cercor/bhs238.

39. Weber LA, Diaconescu AO, Mathys C, Schmidt A, Kometer M, Vollenweider F, Stephan KE. Ketamine Affects Prediction Errors about Statistical Regularities: A Computational Single-Trial Analysis of the Mismatch Negativity. Journal of Neuroscience 2020;40(29):5658–68. URL: https://www.jneurosci.org/content/40/29/5658. doi:10.1523/JNEUROSCI.3069-19.2020; publisher: Society for Neuroscience Section: Research Articles.

40. Timms AE, Dorschner MO, Wechsler J, Choi KY, Kirkwood R, Girirajan S, Baker C, Eichler EE, Korvatska O, Roche KW, Horwitz MS, Tsuang DW. Support for the N-methyl-D-aspartate receptor hypofunction hypothesis of schizophrenia from exome sequencing in multiplex families. JAMA psychiatry 2013;70(6):582– 90. doi:10.1001/jamapsychiatry.2013.1195.

41. Sekar A, Bialas AR, de Rivera H, Davis A, Hammond TR, Kamitaki N, Tooley K, Presumey J, Baum M, Van Doren V, Genovese G, Rose SA, Handsaker RE, Schizophrenia Working Group of the Psychiatric Genomics Consortium, Daly MJ, Carroll MC, Stevens B, McCarroll SA. Schizophrenia risk from complex variation of complement component 4. Nature 2016;530(7589):177–83. doi:10.1038/nature16549.

42. Pocklington AJ, Rees E, Walters JTR, Han J, Kavanagh DH, Chambert KD, Holmans P, Moran JL, McCarroll SA, Kirov G, O’Donovan MC, Owen MJ. Novel Findings from CNVs Implicate Inhibitory and Excitatory Signaling Complexes in Schizophrenia. Neuron 2015;86(5):1203–14. doi:10.1016/j.neuron.2015.04.022.

43. Schizophrenia Working Group of the Psychiatric Genomics Consortium . Biological insights from 108 schizophrenia-associated genetic loci. Nature 2014;511(7510):421–7. doi:10.1038/nature13595.

44. Pers TH, Timshel P, Ripke S, Lent S, Sullivan PF, O’Donovan MC, Franke L, Hirschhorn JN, Schizophrenia Working Group of the Psychiatric Genomics Consortium . Comprehensive analysis of schizophrenia-associated loci highlights ion channel pathways and biologically plausible candidate causal genes. Human Molecular Genetics 2016;25(6):1247–54. doi:10.1093/hmg/ddw007.

45. Gulsuner S, Walsh T, Watts AC, Lee MK, Thornton AM, Casadei S, Rippey C, Shahin H, Nimgaonkar VL, Go RCP, Savage RM, Swerdlow NR, Gur RE, Braff DL, King MC, McClellan JM. Spatial and Temporal Mapping of De novo Mutations in Schizophrenia To a Fetal Prefrontal Cortical Network. Cell 2013;154(3):518–29. URL: https://www.ncbi.nlm.nih.gov/pmc/articles/PMC3894107/. doi:10.1016/j.cell.2013.06.049.

46. Birnbaum R, Jaffe AE, Chen Q, Hyde TM, Kleinman JE, Weinberger DR. Investigation of the Prenatal Expression Patterns of 108 Schizophrenia-Associated Genetic Loci. Biological Psychiatry 2015;77(11):e43–51. URL: https://www.biologicalpsychiatryjournal.com/article/S0006-3223(14)00788-4/abstract. doi:10.1016/j.biopsych.2014.10.008; publisher: Elsevier.

47. Hoftman GD, Datta D, Lewis DA. Layer 3 Excitatory and Inhibitory Circuitry in the Prefrontal Cortex: Developmental Trajectories and Alterations in Schizophrenia. Biological Psychiatry 2017;81(10):862–73. doi:10.1016/j.biopsych.2016.05.022.

48. Wang M, Yang Y, Wang CJ, Gamo NJ, Jin LE, Mazer JA, Morrison JH, Wang XJ, Arnsten AFT. NMDA receptors subserve persistent neuronal firing during working memory in dorsolateral prefrontal cortex. Neuron 2013;77(4):736–49. doi:10.1016/j.neuron.2012.12.032.

49. Coull JT, Morgan H, Cambridge VC, Moore JW, Giorlando F, Adapa R, Corlett PR, Fletcher PC. Ketamine perturbs perception of the flow of time in healthy volunteers. Psychopharmacology 2011;218(3):543– 56. URL: https://doi.org/10.1007/s00213-011-2346-9. doi:10.1007/s00213-011-2346-9.

50. Malhotra AK, Pinals DA, Weingartner H, Sirocco K, Missar CD, Pickar D, Breier A. NMDA receptor function and human cognition: the effects of ketamine in healthy volunteers. Neuropsychopharmacology : official publication of the American College of Neuropsychopharmacology 1996;14:301–7. URL: http://www.ncbi.nlm.nih.gov/pubmed/8703299. doi:ClinicalTrialComparativeStudyControlledClinicalTrial; 5.

51. Moore JW, Turner DC, Corlett PR, Arana FS, Morgan HL, Absalom AR, Adapa R, de Wit S, Everitt JC, Gardner JM, Pigott JS, Haggard P, Fletcher PC. Ketamine administration in healthy volunteers reproduces aberrant agency experiences associated with schizophrenia. Cogn Neuropsychiatry 2011;:1– 18URL: http://www.ncbi.nlm.nih.gov/pubmed/21302161. doi:10.1080/13546805.2010.546074.

52. Lahti AC, Koffel B, LaPorte D, Tamminga CA. Subanesthetic Doses of Ketamine Stimulate Psychosis in Schizophrenia. Neuropsychopharmacology 1995;13(1):9–19. URL: http://www.nature.com/npp/journal/v13/n1/full/1380271a.html. doi:10.1016/0893-133X(94)00131-I.

53. Adams RA, Stephan KE, Brown HR, Frith CD, Friston KJ. The Computational Anatomy of Psychosis. Frontiers in Psychiatry 2013;4. URL: http://www.frontiersin.org/Schizophrenia/10.3389/fpsyt.2013.00047/abstract. doi:10.3389/fpsyt.2013.00047.

54. David O, Friston KJ. A neural mass model for MEG/EEG: coupling and neuronal dynamics. NeuroImage 2003;20:1743–55. URL: http://www.ncbi.nlm.nih.gov/pubmed/14642484. doi:ResearchSupport,Non-U.S.Gov’t; 3.

55. Moran RJ, Kiebel SJ, Stephan KE, Reilly RB, Daunizeau J, Friston KJ. A neural mass model of spectral responses in electrophysiology. NeuroImage 2007;37:706–20. URL: http://www.ncbi.nlm.nih.gov/pubmed/17632015. doi:ResearchSupport,Non-U.S.Gov’t; 3.

56. Jansen BH, Rit VG. Electroencephalogram and visual evoked potential generation in a mathematical model of coupled cortical columns. Biological cybernetics 1995;73:357–66. URL: http://www.ncbi.nlm.nih.gov/pubmed/7578475; x4.

57. Dima D, Frangou S, Burge L, Braeutigam S, James A. Abnormal intrinsic and extrinsic connectivity within the magnetic mismatch negativity brain network in schizophrenia: A preliminary study. Schizophrenia Research 2012;135(1–3):23–7. URL: http://www.sciencedirect.com/science/article/pii/S0920996412000023. doi:10.1016/j.schres.2011.12.024.

58. Dima D, Dietrich DE, Dillo W, Emrich HM. Impaired top-down processes in schizophrenia: A DCM study of ERPs. NeuroImage 2010;52(3):824–32. URL: http://www.sciencedirect.com/science/article/pii/S1053811909013755. doi:10.1016/j.neuroimage.2009.12.086.

59. Dima D, Roiser JP, Dietrich DE, Bonnemann C, Lanfermann H, Emrich HM, Dillo W. Understanding why patients with schizophrenia do not perceive the hollow-mask illusion using dynamic causal modelling. NeuroImage 2009;46(4):1180–6. URL: http://www.sciencedirect.com/science/article/pii/S105381190900278X. doi:10.1016/j.neuroimage.2009.03.033.

60. Fogelson N, Litvak V, Peled A, Fernandez-del Olmo M, Friston K. The functional anatomy of schizophrenia: A dynamic causal modeling study of predictive coding. Schizophrenia Research 2014;158(1):204– 12. URL: http://www.sciencedirect.com/science/article/pii/S0920996414003028. doi:10.1016/j.schres.2014.06.011.

61. Nelson HE. THE NATIONAL ADULT READING TEST (NART) ????;.

62. McGlashan TH, Johannessen JO. Early detection and intervention with schizophrenia: rationale. Schizophrenia Bulletin 1996;22(2):201–22. doi:10.1093/schbul/22.2.201.

63. Schmidt A, Cappucciati M, Radua J, Rutigliano G, Rocchetti M, Dell’Osso L, Politi P, Borgwardt S, Reilly T, Valmaggia L, McGuire P, Fusar-Poli P. Improving Prognostic Accuracy in Subjects at Clinical High Risk for Psychosis: Systematic Review of Predictive Models and Meta-analytical Sequential Testing Simulation. Schizophrenia Bulletin 2017;43(2):375–88. doi:10.1093/schbul/sbw098.

64. Kiang M, Patriciu I, Roy C, Christensen BK, Zipursky RB. Test-retest reliability and stability of N400 effects in a word-pair semantic priming paradigm. Clinical Neurophysiology: Official Journal of the International Federation of Clinical Neurophysiology 2013;124(4):667–74. doi:10.1016/j.clinph.2012.09.029.

65. Nelson DL, McEvoy CL, Schreiber TA. The University of South Florida free association, rhyme, and word fragment norms. Behavior Research Methods, Instruments, & Computers 2004;36(3):402–7. URL: https://doi.org/10.3758/BF03195588. doi:10.3758/BF03195588.

66. Litvak V, Mattout J, Kiebel S, Phillips C, Henson R, Kilner J, Barnes G, Oostenveld R, Daunizeau J, Flandin G, Penny W, Friston K. EEG and MEG Data Analysis in SPM8. Computational Intelligence and Neuroscience 2011;2011:1–32. URL: http://www.hindawi.com/journals/cin/2011/852961/. doi:10.1155/2011/852961.

67. Momi D, Wang Z, Griffiths JD. TMS-evoked responses are driven by recurrent large-scale network dynamics. eLife 2023;12:e83232. URL: https://doi.org/10.7554/eLife.83232. doi:10.7554/eLife.83232; publisher: eLife Sciences Publications, Ltd.

68. Mozer M. A Focused Backpropagation Algorithm for Temporal Pattern Recognition. Complex Syst 1989;URL: https://www.semanticscholar.org/paper/A-Focused-Backpropagation-Algorithm-for-Temporal-Mozer/7fb4d10f6d2ee3133135958aefd50bf22dcced9d.

69. Robinson AJ, Fallside F. Static and dynamic error propagation networks with application to speech coding. In: Proceedings of the 1st International Conference on Neural Information Processing Systems. NIPS’87; Cambridge, MA, USA: MIT Press; 1987:632–41.

70. Werbos PJ. Generalization of backpropagation with application to a recurrent gas market model. Neural Networks 1988;1(4):339–56. URL: https://www.sciencedirect.com/science/article/pii/089360808890007X. doi:10.1016/0893-6080(88)90007-X.

71. Schaefer A, Kong R, Gordon EM, Laumann TO, Zuo XN, Holmes AJ, Eickhoff SB, Yeo BTT. Local-Global Parcellation of the Human Cerebral Cortex from Intrinsic Functional Connectivity MRI. Cerebral Cortex (New York, NY: 1991) 2018;28(9):3095–114. doi:10.1093/cercor/bhx179.

72. Yeo BTT, Krienen FM, Sepulcre J, Sabuncu MR, Lashkari D, Hollinshead M, Roffman JL, Smoller JW, Zöllei L, Polimeni JR, Fischl B, Liu H, Buckner RL. The organization of the human cerebral cortex estimated by intrinsic functional connectivity. Journal of Neurophysiology 2011;106(3):1125–65. doi:10.1152/jn.00338.2011.

73. Krishnan A, Williams LJ, McIntosh AR, Abdi H. Partial Least Squares (PLS) methods for neuroimaging: A tutorial and review. NeuroImage 2011;56(2):455–75. URL: http://linkinghub.elsevier.com/retrieve/pii/S1053811910010074. doi:10.1016/j.neuroimage.2010.07.034.

74. Wang XJ, Krystal J. Computational Psychiatry. Neuron 2014;84(3):638–54. URL: http://www.sciencedirect.com/science/article/pii/S0896627314009167. doi:10.1016/j.neuron.2014.10.018.

75. Jackson F, Foti D, Kotov R, Perlman G, Mathalon DH, Proudfit GH. An incongruent reality: The N400 in relation to psychosis and recovery. Schizophrenia Research 2014;160(1):208–15. URL: https://www.sciencedirect.com/science/article/pii/S0920996414005313. doi:10.1016/j.schres.2014.09.039.

76. Perrottelli A, Giordano GM, Brando F, Giuliani L, Mucci A. EEG-Based Measures in At-Risk Mental State and Early Stages of Schizophrenia: A Systematic Review. Frontiers in Psychiatry 2021;12. URL: https://www.frontiersin.org/journals/psychiatry/articles/10.3389/fpsyt.2021.653642/full. doi:10.3389/fpsyt.2021.653642; publisher: Frontiers.

77. Wang K, Cheung EFC, Gong Qy, Chan RCK. Semantic Processing Disturbance in Patients with Schizophrenia: A Meta-Analysis of the N400 Component. PLOS ONE 2011;6(10):e25435. URL: https://journals.plos.org/plosone/article?id=10.1371/journal.pone.0025435. doi:10.1371/journal.pone.0025435; publisher: Public Library of Science.

78. Lepock JR, Ahmed S, Mizrahi R, Gerritsen CJ, Maheandiran M, Bagby RM, Korostil M, Kiang M. N400 event-related brain potential as an index of real-world and neurocognitive function in patients at clinical high risk for schizophrenia. Early Intervention in Psychiatry 2021;15(1):68–75. doi:10.1111/eip.12911.

79. Friston K. A theory of cortical responses. Philosophical transactions of the Royal Society of London Series B, Biological sciences 2005;360:815–36. URL: http://www.ncbi.nlm.nih.gov/pubmed/15937014. doi:ResearchSupport,Non-U.S.Gov’tReview; 1456.

80. Servan-Schreiber D, Cohen JD, Steingard S. Schizophrenic deficits in the processing of context. A test of a theoretical model. Archives of general psychiatry 1996;53:1105–12. URL: http://www.ncbi.nlm.nih.gov/pubmed/8956676. doi:ResearchSupport,U.S.Gov’t,P.H.S.; 12.

81. MacDonald AW, Carter CS, Kerns JG, Ursu S, Barch DM, Holmes AJ, Stenger VA, Cohen JD. Specificity of Prefrontal Dysfunction and Context Processing Deficits to Schizophrenia in Never-Medicated Patients With First-Episode Psychosis. American Journal of Psychiatry 2005;162(3):475–84. URL: http://ajp.psychiatryonline.org/doi/abs/10.1176/appi.ajp.162.3.475. doi:10.1176/appi.ajp.162.3.475.

82. Bastos AM, Usrey WM, Adams RA, Mangun GR, Fries P, Friston KJ. Canonical microcircuits for predictive coding. Neuron 2012;76(4):695–711. URL: https://www.ncbi.nlm.nih.gov/pmc/articles/PMC3777738/. doi:10.1016/j.neuron.2012.10.038.

83. Bastos-Leite AJ, Ridgway GR, Silveira C, Norton A, Reis S, Friston KJ. Dysconnectivity Within the Default Mode in First-Episode Schizophrenia: A Stochastic Dynamic Causal Modeling Study With Functional Magnetic Resonance Imaging. Schizophrenia Bulletin 2015;41(1):144–53. URL: http://www.ncbi.nlm.nih.gov/pmc/articles/PMC4266292/. doi:10.1093/schbul/sbu080.

84. Sterzer P, Adams RA, Fletcher P, Frith C, Lawrie SM, Muckli L, Petrovic P, Uhlhaas P, Voss M, Corlett PR. The predictive coding account of psychosis. Biological Psychiatry 2018;URL: https://www.sciencedirect.com/science/article/pii/S0006322318315324. doi:10.1016/j.biopsych.2018.05.015.

85. Teufel C, Subramaniam N, Dobler V, Perez J, Finnemann J, Mehta PR, Goodyer IM, Fletcher PC. Shift toward prior knowledge confers a perceptual advantage in early psychosis and psychosis-prone healthy individuals. Proceedings of the National Academy of Sciences 2015;112(43):13401–6. URL: http://www.pnas.org/content/112/43/13401. doi:10.1073/pnas.1503916112.

86. Adams RA, Huys QJM, Roiser JP. Computational Psychiatry: towards a mathematically informed understanding of mental illness. Journal of Neurology, Neurosurgery & Psychiatry 2015;:jnnp–2015URL: http://jnnp.bmj.com/content/early/2015/07/08/jnnp-2015-310737. doi:10.1136/jnnp-2015-310737.

87. Fryer S, Roach B, Hamilton H, Bachman P, Belger A, Carrion R, Duncan E, Johannesen J, Light G, Niznikiewicz M, Addington J, Bearden C, Cadenhead K, Cannon TD, Cornblatt B, McGlashan T, Perkins D, Seidman L, Tsuang M, Walker E, Woods S, Mathalon D. Deficits in Auditory Predictive Coding in Individuals With the Psychosis Risk Syndrome: Prediction of Conversion to Psychosis. Biological Psychiatry 2020;87(9):S117–8. URL: https://www.biologicalpsychiatryjournal.com/article/S0006-3223(20)30433-9/abstract. doi:10.1016/j.biopsych.2020.02.321; publisher: Elsevier.

88. Charlton CE, Lepock JR, Hauke DJ, Mizrahi R, Kiang M, Diaconescu AO. Atypical prediction error learning is associated with prodromal symptoms in individuals at clinical high risk for psychosis. Schizophrenia 2022;8(1):1–10. URL: https://www.nature.com/articles/s41537-022-00302-3. doi:10.1038/s41537-022-00302-3; number: 1 Publisher: Nature Publishing Group.

89. Hauke DJ, Charlton CE, Schmidt A, Griffiths JD, Woods SW, Ford JM, Srihari VH, Roth V, Diaconescu AO, Mathalon DH. Aberrant Hierarchical Prediction Errors Are Associated With Transition to Psychosis: A Computational Single-Trial Analysis of the Mismatch Negativity. Biological Psychiatry: Cognitive Neuroscience and Neuroimaging 2023;8(12):1176–85. URL: https://www.sciencedirect.com/science/article/pii/S2451902223001921. doi:10.1016/j.bpsc.2023.07.011.

90. Hamilton HK, Roach BJ, Bachman PM, Belger A, Carrión RE, Duncan E, Johannesen JK, Light GA, Niznikiewicz MA, Addington J, Bearden CE, Cadenhead KS, Cornblatt BA, McGlashan TH, Perkins DO, Tsuang MT, Walker EF, Woods SW, Cannon TD, Mathalon DH. Mismatch Negativity in Response to Auditory Deviance and Risk for Future Psychosis in Youth at Clinical High Risk for Psychosis. JAMA Psychiatry 2022;79(8):780–9. URL: https://doi.org/10.1001/jamapsychiatry.2022.1417. doi:10.1001/jamapsychiatry.2022.1417.

91. Das T, Borgwardt S, Hauke DJ, Harrisberger F, Lang UE, Riecher-Rössler A, Palaniyappan L, Schmidt Disorganized Gyrification Network Properties During the Transition to Psychosis. JAMA Psychiatry 2018;75(6):613–22. URL: https://jamanetwork.com/journals/jamapsychiatry/fullarticle/2678576. doi:10.1001/jamapsychiatry.2018.0391; publisher: American Medical Association.

